# Impact of the COVID-19 Pandemic on Same-day Discharge Adoption Following Percutaneous Coronary Intervention for Stable CAD: A National Time Series Analysis

**DOI:** 10.1101/2023.08.28.23294753

**Authors:** Manoj Thangam, Michael Grzeskowiak, Frederick A Masoudi, Matthew C Bunte, Edward Fry, Mark Pirwitz, Allison Ottenbacher, Ana Cristina Perez, Collin Miller, Sarah Benedict, Amit Amin, Sahil A. Parikh, Peter Monteleone

**Author notes:** Corresponding Author: Manoj Thangam MD, Ascension Texas Cardiovascular, The University of Texas at Austin Dell School of Medicine, 1301 West 38^th^ Street, Suite 400, Austin, Texas 78701.

## Abstract

**Background:** Elective percutaneous coronary intervention (PCI) historically required hospitalization post procedure. Same day discharge (SDD) has emerged as a safe and cost efficient option, although the impact of the coronavirus disease of 2019 (COVID-19) pandemic on rates of SDD and associated care episode costs remains uncertain.

**Methods:** A national sample of consecutive patients undergoing elective PCI at 42 hospitals (Ascension, St.Louis, MO) between May 2019 to April 2021 were identified using internal registry data and administrative claims data. Rates of SDD before and after the COVID-19 pandemic (March 2020) were compared using multivariable logistic regression adjusted for patient and procedural characteristics. Additionally, an interrupted time series model was used to determine the effect of the pandemic and policy on SDD rates before and after pandemic declaration. Lastly, we estimated total costs per PCI episode in pre and post pandemic periods.

**Results:** In total, 12,740 interventions were performed within 42 Ascension facilities that met study eligibility criteria (5955 PCI prior to the pandemic and 6785 after). Demographic data were similar between both populations although higher rates of dyslipidemia, prior myocardial infarction, and heart failure history were noted in the post pandemic group. Pandemic declaration was associated with a higher likelihood of SDD (OR 2.09, CI 1.93-2.25, p < 0.001). From pre-pandemic to post-pandemic, mean SDD rose from 34% to 45% (p< 0.001) with an accelerated monthly SDD adoption rate after the pandemic (0.1% per month vs 1.0% per month, p=0.02). Total costs per episode were $679.52 (95% CI $476.12 – $882.92, p < 0.001) higher in the post-pandemic period, driven by increased material costs. SDD was associated with a $2137.05 (95% CI $1925.03 – $2349.07, p < 0.001) reduction in costs relative to non-SDD episodes throughout the study period.

**Conclusion:** Among a large national risk-adjusted sample of consecutive patients, the COVID-19 pandemic accelerated adoption of SDD. As a care strategy, SDD was associated with reduced episode costs during elective PCI in the post-pandemic period.

## Introduction

Percutaneous coronary intervention (PCI) remains a commonly used therapy to treat symptomatic obstructive coronary artery disease (CAD) with about 600,000 interventions performed annually in the US.^1,2,3^ With the evolution of PCI, procedural efficacy and safety have improved considerably.^4,5^ During the early period of coronary intervention, patients were often observed in the hospital for days due to concern for acute stent thrombosis, coronary dissection, procedural failure, antithrombotic regimens, and access site complications.^5^ Substantial improvements in procedural techniques, technology, and operator experience have facilitated safe same day discharge (SDD) among selected populations undergoing elective PCI.^4,5,6^ Several studies have demonstrated the safety of SDD following elective PCI for stable CAD within regional centers and national registries,^7–12^ with no difference in 30-day all-cause mortality or rehospitalizations.^11^ Furthermore, SDD is associated with no difference in access site complications or acute kidney injury.^13,14^ The evidence base supporting SDD utilization has resulted in guideline recommendations encouraging adoption of SDD in suitable patient populations undergoing elective PCI.^14,15^

The increasing costs of cardiovascular care highlights the importance of judicious resource allocation and the need to tailor healthcare policies to improve costs concurrent with quality of care.^16^ Policy measures including the Affordable Care Act and ensuing bundled payment programs have incentivized the delivery of higher value care by enhancing quality, reducing cost, or both.^17^ In the United States, SDD has been estimated to save about $5000 per procedure with a potential national cost savings approaching $577 million through widespread adoption with savings driven primarily by reduced supply utilization along with room and board expenditures.^7^ In addition to the cost savings inherent to reducing length of stay, the value of bed availability is more pertinent than ever before considering the COVID-19 global pandemic. This pandemic placed a spotlight on the importance of reducing hospital admissions and improving cost within our strained health system. Ascension organized a collaborative system wide effort focusing on SDD rates to meet these demands. The potential for improving cost, efficiency, and bed availability using SDD without compromising patient outcomes during the COVD-19 pandemic prompted the development and implementation of our SDD directive at Ascension, a large US health system.

Accordingly, the objectives of this analysis were to describe characteristics among those allocated to SDD after elective PCI for stable CAD, effectiveness of an institutional SDD implementation, as well as describe trends relative to the onset of the COVID-19 pandemic in SDD utilization and the impact of SDD on total care costs. These results may expose opportunities to improve appropriate adoption of SDD and thereby enhance effectiveness and efficiency of PCI for stable CAD.

## Methods

### Study Design and Participants

This was a retrospective observational analysis from a large non-profit hospital network (Ascension, St. Louis, MO) of 151 hospitals in 21 states and the District of Columbia evaluating the impact of the COVID-19 pandemic and a national quality improvement program on SDD utilization using a time series design. Patients were identified by evaluation of the Ascension internal National Cardiovascular Data Registry (NCDR) CathPCI registry. Consecutive adults aged > 18 years undergoing elective PCI with survival to hospital discharge treated between May 2019 and April 2021 were eligible for inclusion. The study was reviewed and approved by the central institutional review board with informed consent waived as a quality improvement initiative.

### Variables

The primary outcome was SDD rate, which was compared before and after declaration of the COVID-19 pandemic (March 2020). Secondary endpoints included the SDD rate after implementation of a national SDD program (May 2020) and total episode costs to evaluate the impact of SDD utilization. Total episode costs were normalized across the study period and estimated using institutional accounting software that provided nationalized average cost for PCI encounter within the Ascension Health System. Subjects that were ineligible for SDD were also excluded from the analysis and included those with the following concurrent conditions: cardiogenic shock, major procedural complications including perforation and dissection, bleeding or vascular complication, PCI indication of acute coronary syndrome (ST elevation myocardial infarction, non-ST elevation myocardial infarction, and unstable angina), use of mechanical circulatory support (intra-aortic balloon pump, implantable intracardiac pump, or extracorporeal membrane oxygenation), cardiac arrest during or prior to admission, post procedural heart failure, cardiac tamponade, or any post procedure stroke. The definitions for exclusion criteria (cardiogenic shock, procedural complications, acute coronary syndrome, mechanical circulatory support, cardiac arrest, heart failure, cardiac tamponade, and stroke) are aligned with those established by the NCDR Cath PCI Registry.

### Data

Clinical data were collected and aggregated from internal system wide NCDR Cath-PCI registry entries using proprietary quality data abstraction software and merged with internal billing data. In addition to conforming to the requirements of the ACC-NCDR data quality program^18^, including periodic external audits, Ascension performs weekly data quality audits to ensure the consistency and accuracy of data abstraction. Patients identified from this registry were linked to cost accounting data for our analysis. Each encounter is divided into cost categories and the aggregate sum of all categories for admission encompasses the total encounter cost. To account for variability in cost over the study periods and across markets, costs were normalized by deriving an average cost per item to facilitate standardization despite time and location of hospital encounter.

### Same Day Discharge Policy

Prior to the COVID-19 pandemic, SDD occurred within individual Ascension hospitals and operators without specific system-wide clinical guidance. With the operational impact of the pandemic, clinicians were confronted with the need to reduce patient and staff infection exposure risk, costs associated with overnight admission, and optimize PCI efficiencies. In response to these needs, a committee of interventional cardiologists (Ascension Interventional Cardiology Affinity Group) representing several of its PCI programs developed a SDD protocol that was approved and implemented in May 2020 (see Supplemental Figure 1).

The SDD policy implementation that began in May 2020 was unique in several key aspects. First, individual hospitals were offered organizational support in the form of shared policy templates, educational resources, and clinical guidance. However, each was allowed to develop and execute individualized hospital policies to increase SDD. Second, each site was informed that their SDD rates would be tracked with a minimum goal of increasing SDD among elective PCIs to ≥ 15% of total PCIs performed (the ACC NCDR Cath PCI average in March 2020).

Additionally, individual center SDD rates would be assessed in the context of rates nationally and among Ascension sites. Lastly, minimal financial expenditures were required in the development, initiation, and monitoring of this policy since each individual center was responsible for policy development and SDD monitoring was integrated into established quality metrics that are evaluated on an ongoing basis.

The COVID-19 pandemic affected elective PCI volumes, delivery, and availability of hospital beds. Therefore, evaluating the impact of the SDD initiative separate from the pandemic effect was not plausible. To best represent the data, we evaluated SDD among the elective PCI population prior and post declaration of the pandemic (March 2020). Notably, the implementation of the Ascension SDD policy did not occur until May 2020 following the national pandemic declaration. Therefore, the study focused on SDD rates pre and post pandemic understanding that the SDD policy also effected SDD adoption throughout our medical system.

### Statistics

Demographic characteristics are presented as mean ± SD for continuous variables and proportions for categorical variables. Demographics between those treated prior to the pandemic were compared to those treated post using either the Pearsons chi squared test or Student’s T-test as appropriate. We evaluated the likelihood of SDD using a logistic regression model adjusting for significant confounders between the pre-pandemic and post-pandemic population. Confounders included age, sex, Black race, treatment period with reference to the pandemic, heart failure, and radial access. We performed a segmented regression with a monthly interrupted time series to better quantify the expected number of SDD before and after the pandemic. A linear relationship between time and average monthly SDD was assumed, and we used New-West standard errors for coefficients estimated by ordinary least-squares regression. Lastly, we evaluated the cost associated with elective PCI with a general linear regression model adjusting for the pandemic timeframe (before and after declaration March 2020), SDD, and additional covariates. All covariates for the different models were identified by univariate significance or were historically known significant variables. All statistical analyses were performed using R statistical software version 4.0.2 (R Project for Statistical Computing) and Stata version 17 (Stata Corp, College Station, Texas). A 2-sided p value of ≤ 0.05 was considered statistically significant.

## Results

Between May 2019 to April 2021, 12,740 patients undergoing PCI for stable CAD were analyzed, including those receiving SDD (n=5588) and non-SDD (n=7152). Patients discharged the same day tended to be younger (67.0 vs 68.9, p<0.001), less frequently female (27.2% vs 32.5%, p<0.001) and Black (5.9% vs 8.0%, p<0.001) (see Supplemental Table 1). Those receiving SDD were less likely to have comorbidities including peripheral artery disease (15.6% vs 17.9%, p<0.001), diabetes mellitus (43.6% vs 46.2%, p<0.001), receive dialysis (1.5% vs 2.5%, p<0.001), and have heart failure (22.6% vs 29.2%, p<0.001). Access site for PCI was also significantly different among discharge cohorts (ANOVA p<0.001), with radial access being utilized more frequently in the SDD cohort (68.9% vs 41.7%, p < 0.001).

Among those receiving SDD, age, sex, the proportion of Black and Hispanic patients and other clinical characteristics were similar between pre and post pandemic groups (see Table 1). However, those receiving elective PCI after the pandemic had a higher percentage of dyslipidemia (p<0.001), prior myocardial infarction (p<0.001), and history of heart failure (p<0.001). Access site for PCI did not differ between SDD groups before and after onset of the pandemic.

**Table 1:** Baseline characteristics among those undergoing SDD after elective PCI, according to period of treatment before or following onset of the COVID-19 pandemic.

| Baseline characteristic | SDD Before Pandemic<br>(May 2019 – April 2020)<br>N= 5955 | SDD After Pandemic<br>(March 2020 – April 2021)<br>N= 6785 | P value |
| --- | --- | --- | --- |
| Age, mean (SD) | 68.1 (10.5) | 68.0 (10.4) | 0.69 |
| Female | 1819 (30.5%) | 2024 (29.8%) | 0.38 |
| Black\ African American | 418 (7.0%) | 481 (7.1%) | 0.88 |
| Hispanic | 177 (3.0%) | 169 (2.5%) | 0.10 |
| Hypertension | 5584 (93.8%) | 6357 (93.7%) | 0.86 |
| Dyslipidemia | 5451 (91.5%) | 6390 (94.2%) | <0.001 |
| Prior MI | 1929 (32.4%) | 2362 (34.8%) | <0.001 |
| PAD | 985 (16.5%) | 1167 (17.2%) | 0.33 |
| Prior PCI | 3232 (54.3%) | 3703 (54.6%) | 0.73 |
| DM | 2680 (45.0%) | 3065 (45.2%) | 0.85 |
| Currently on Dialysis | 131 (2.2%) | 136 (2.0%) | 0.44 |
| HF | 1457 (24.5%) | 1892 (27.9%) | <0.001 |
| Tobacco Use |  |  |  |
| Current | 1056 (17.7%) | 1230 (18.1%) | 0.33 |
| Former | 2510 (42.1%) | 2837 (41.8%) |  |
| Never | 2264 (38.0%) | 2604 (38.4%) |  |
| Unknown | 125 (2.1%) | 114 (1.7%) |  |
| Access site |  |  | 0.47 |
| Femoral | 2772 (46.5%) | 3085 (45.5%) |  |
| Radial | 3158 (53.0%) | 3670 (54.1%) |  |
| Other | 25 (0.4%) | 30 (0.4%) |  |

IQR: interquartile range, SD: standard deviation, MI: myocardial infarction, PAD: peripheral
artery disease, PCI: percutaneous coronary intervention, DM: diabetes mellitus, HF: heart
failure, SDD: same day discharge.

Nationally, the rate of SDD increased significantly after the onset of the pandemic (34% vs 45%, p=< 0.001) (see Figure 1). In segmented monthly interrupted time series analysis, the mean month-over-month rate of SDD adoption increased by 0.1% and 1.0% in pre-pandemic and post-pandemic periods, respectively (p= 0.02) (see Figure 2). The odds of SDD were higher among men (1.23, 1.13-1.33, p<0.001), non-Black patients (1.39, 1.20-1.62, p<0.001), those treated in the post-pandemic period (OR 2.09, CI 1.93-2.25, p < 0.001), and among those receiving radial access (OR 3.05, 2.83-3.29, p < 0.001) (see Table 2).The odds of SDD were lower with increasing age (OR 0.99, 0.98-0.99, p<0.001) and among those with heart failure (0.77, 0.71-0.84, p<0.001).

**Figure 1:**
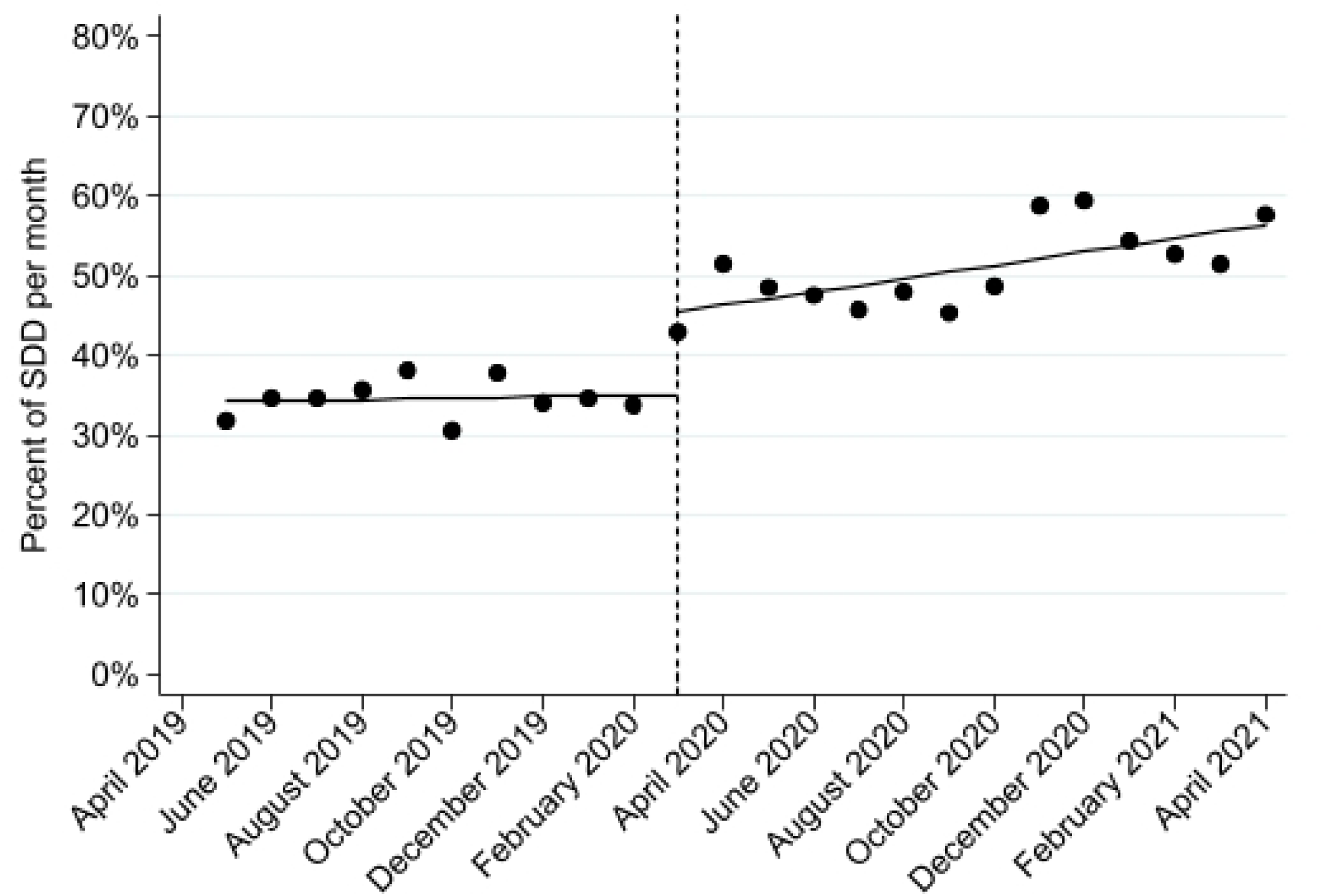
Segmented Regression with Interrupted Time Series Analysis Displaying Pre Policy and Post Policy Same Day Discharge Percentages.

**Table 2:** Predictors of SDD during PCI for stable CAD.

| Baseline characteristic | Odds<br>Ratio | 95% CI | P-value |
| --- | --- | --- | --- |
| Age | 0.99 | 0.98 to 0.99 | <0.001 |
| Male | 1.23 | 1.13 to 1.33 | <0.001 |
| After Pandemic | 2.09 | 1.94 to 2.26 | <0.001 |
| Non-African American | 1.39 | 1.20 to 1.62 | <0.001 |
| Heart Failure | 0.77 | 0.71 to 0.84 | <0.001 |
| Radial Access | 3.05 | 2.83 to 3.29 | <0.001 |

CI: confidence interval, Age was evaluated with 1 year increase regarding coefficients and odds
ratio. PCI: percutaneous coronary intervention, HF: heart failure.

By episode of care, SDD was associated with significant total cost savings of $2137.05 (95% CI $1925.02 to $2349.07 less then pre-pandemic, p < 0.001) per PCI hospital encounter after accounting for inter year cost variation of pre and post pandemic and other relevant covariates. The cost variability pre and post pandemic was noted to be an average increase in material cost of $679.52 (95% CI $476.12 to $882.92; p value < 0.001) in the post pandemic period.

Several variables were associated with increased total episode costs (see Table 3). The greatest drivers of increased costs included heart failure ($1167.15 per episode, 95% CI, $936.93-$1398.08; p<0.001) and peripheral artery disease ($713.91 per episode, 95% CI, $443.14-$984.68). The factors associated with the largest reductions in episodic care costs following PCI for stable CAD included SDD (-$2731.05 per episode, 95% CI, –$2349.08-–$1925.03; p<0.001) and non-Black race (-$910.77 per episode, 95% CI, –$1302.68 – –$518.87;p<0.001).

**Table 3:** Risk-adjusted contributors to total episodic care costs (USD) during PCI for stable CAD.

| Baseline characteristic | Coefficient | 95% CI | P value |
| --- | --- | --- | --- |
| Age* | \$24.60 | \$14.45 to \$34.74 | <0.001 |
| Male | \$379.91 | \$11.96 to \$598.12 | 0.001 |
| After Pandemic | \$679.5 | \$476.12 to \$882.92 | <0.001 |
| Same day Discharge | -\$2731.05 | -\$2349.07 to -<br>\$1925.03 | <0.001 |
| Non-African American | -\$910.77 | -\$1302.68 to -\$518.87 | <0.001 |
| Peripheral artery disease | \$713.91 | \$443.14 to \$ 984.68 | <0.001 |
| HF | \$1167.15 | \$936.93 to \$1398.08 | <0.001 |
| Prior PCI | -\$828.90 | -\$1034.06 to -\$623.74 | <0.001 |
| Dyslipidemia | -\$541.48 | -\$937.93 to -\$145.02 | <0.01 |
| Diabetes | \$285.53 | \$81.99 to \$489.06 | <0.001 |
| Current Smoker | \$427.89 | \$153.99 to \$701.80 | <0.01 |
| Radial access | -\$484.34 | -\$694.38 to -\$274.29 | <0.001 |

\* Per one year increase in age, CI: confidence interval, PCI: percutaneous coronary intervention,
HF: heart failure.

## Discussion

In this large, national, consecutive experience of SDD before and after the COVID-19 pandemic we found SDD rates increased over the study period. The increase in SDD rates persisted along with a trend in monthly growth of SDD rates at 1 year after onset of the pandemic. The adoption of SDD was associated with reduced total episode costs despite a relative increase in overall costs of elective PCI in the post pandemic period.

The SDD protocol emerged from ongoing quality improvement efforts as well as in response to the COVID –19 pandemic within the Ascension Interventional Cardiology Affinity Group. Before the pandemic and implementation of the SDD protocol, wide variation of SDD was noted among individual centers, with individual institutional rates below the national registry average for many. Initiation of this protocol established SDD rates as a quality metric actively tracked by our health system with the intention to hold individual centers and operators accountable. The post pandemic period along with the SDD protocol facilitated in improvement in SDD rates throughout Ascension Healthcare. Although aggregate trends across the system showed an increase in SDD, there was variability among individual centers with a minority showing no change or a decrease in SDD. These were primarily small centers with low PCI volumes and likely represents variability inherent to low volume centers.

The use of radial access has been previously reported to be associated with reduced periprocedural bleeding and reduced length of stay.^6,7,19,20^ This analysis found that radial access had the strongest influence on a SDD strategy, though rates of radial access use were not significantly different before and after the onset of the pandemic. Therefore, the adoption of radial access for PCI of stable CAD may contribute to the success of a SDD strategy and facilitate the other tangible benefits of reduced healthcare utilization also observed in this analysis.

Our study showed that SDD facilitated cost savings during the post pandemic period within our health system. The increase in cost after onset of the pandemic is attributed to the increase in supplies and facilities due to inflation over the study period. Despite this increase in cost of elective PCI after the pandemic declaration, there was significant reduction in total episodic costs. This highlights the potential for SDD to facilitate improved healthcare efficiency even with increasing equipment and facility costs currently experienced by systems throughout the country.

Another important aspect of the SDD policy implementation was its potential to improve hospital bed availability. The COVID-19 pandemic impacted health care systems throughout the world. With the CDC declaration of the pandemic in March 2020, the SDD policy seemed more relevant than ever before to facilitate needed procedures in a world where hospital occupancy was pushed to the brink.^21,22^ Our SDD policy was developed over a 6-month period prior to enactment in May of 2020; however, the COVID-19 pandemic effected the implementation and adoption of the policy. It is likely that the policy improvement of SDD was impacted by the lack of available beds during the pandemic and the desire to reduce hospital stays as much as possible. However, the improvement in SDD has been persistent in our national system at follow up extending to 12 months after pandemic declaration supporting some persistent impact from the SDD policy.

## Study Limitations

Several limitations are inherent to this retrospective observational analysis and deserve mention. First, the effect of the COVID-19 pandemic on SDD policy adoption cannot be attributed based on the nature of this evaluation being non-randomized and which may be confounded by contemporaneous institutional efforts to promote SDD. The effect of COVID on the SDD policy likely increased SDD rates and policy adoption. Yet, the persistent increase in SDD supports the continued effect of the policy on improving SDD among patients undergoing PCI for stable CAD. Second, although Ascension is a large nationally represented health system the generalizability of these results are uncertain and require additional confirmation.

Furthermore, female patients, non-White, and non-Hispanic patients were underrepresented relative to estimates within the national population Third, unidentified confounders, including characteristics of the medical history and prior treatment, as well as circumstances of the population, including health-related social needs, were not measured. Future research should explore the utility of SDD among the underserved community to confirm consistent benefits.

We included a consecutive population to limit selection bias among those undergoing PCI for stable CAD. Lastly, the SDD policy developed by individual centers had minor variability due to the discretion of individual centers. Therefore, despite a national policy mandate, individual policies and its implementation varied between Ascension sites. This study did not evaluate post-discharge outcomes, including rates of major adverse cardiovascular events or rates of readmission, to assess safety of the SDD strategy since post discharge data were unavailable for evaluation. Furthermore, prior studies have shown SDD to be safe in selected PCI populations.^4,5,6^

## Conclusion

From a large consecutive national sample, rates of SDD increased following the onset of the COVID-19 pandemic and were subsequently associated with concurrent reductions in episode care costs. Initiatives that promote SDD and radial access may facilitate expansion of SDD adoption and thereby extend the clinical benefits and efficiencies provided by both care strategies.

## Data Availability

All data used in this analysis are available in our secured system for clarification and additional analysis as needed.

## Acknowledgements

None

## Supplemental Appendix

**Table 1:** Baseline characteristics of patients undergoing elective PCI, according to same-day discharge status.

| Baseline characteristic | Non-SDD<br>N=7152 | SDD<br>N=5588 | P value |
| --- | --- | --- | --- |
| Age, mean (SD) | 68.9 (10.4) | 67.0 (10.3) | <0.001 |
| Female | 2323 (32.5%) | 1520 (27.2%) | <0.001 |
| Black\ African American | 572 (8.0%) | 327 (5.9%) | <0.001 |
| Hispanic | 197 (2.8%) | 149 (2.7%) | 0.80 |
| Hypertension | 6706 (93.8%) | 5235 (93.7%) | 0.85 |
| Dyslipidemia | 6621 (92.6%) | 5220 (93.4%) | 0.07 |
| Prior MI | 2393 (33.5%) | 1898 (34.0%) | 0.55 |
| PAD (Peripheral Artery Disease) | 1279 (17.9%) | 873 (15.6%) | <0.001 |
| Prior PCI | 3840 (53.7%) | 3095 (55.4%) | 0.06 |
| DM | 3307 (46.2%) | 2438 (43.6%) | <0.01 |
| Currently on Dialysis | 181 (2.5%) | 86 (1.5%) | <0.001 |
| HF | 2088 (29.2%) | 1261 (22.6%) | <0.001 |
| Tobacco Use |  |  |  |
| Current | 1207 (16.9%) | 1079 (19.3%) | <0.001 |
| Former | 3092 (43.2%) | 2255 (40.4%) |  |
| Never | 2715 (38.0%) | 2153 (38.5%) |  |
| Unknown | 138 (1.9%) | 101 (1.8%) |  |
| Access site |  |  | <0.001 |
| Femoral | 4141 (57.9%) | 1716 (30.7%) |  |
| Radial | 2979 (41.7%) | 3849 (68.9%) |  |
| Other | 32 (0.4%) | 23 (0.4%) |  |
| Pandemic |  |  | <0.001 |
| Before | 3865 (54.0%) | 2090 (37.4%) |  |
| After | 3287 (46.0%) | 3498 (62.6%) |  |

IQR: interquartile range, SD: standard deviation, MI: myocardial infarction, PAD: peripheral
artery disease, PCI: percutaneous coronary intervention, DM: diabetes mellitus, HF: heart
failure, SDD: same day discharge.

**Figure 1:**
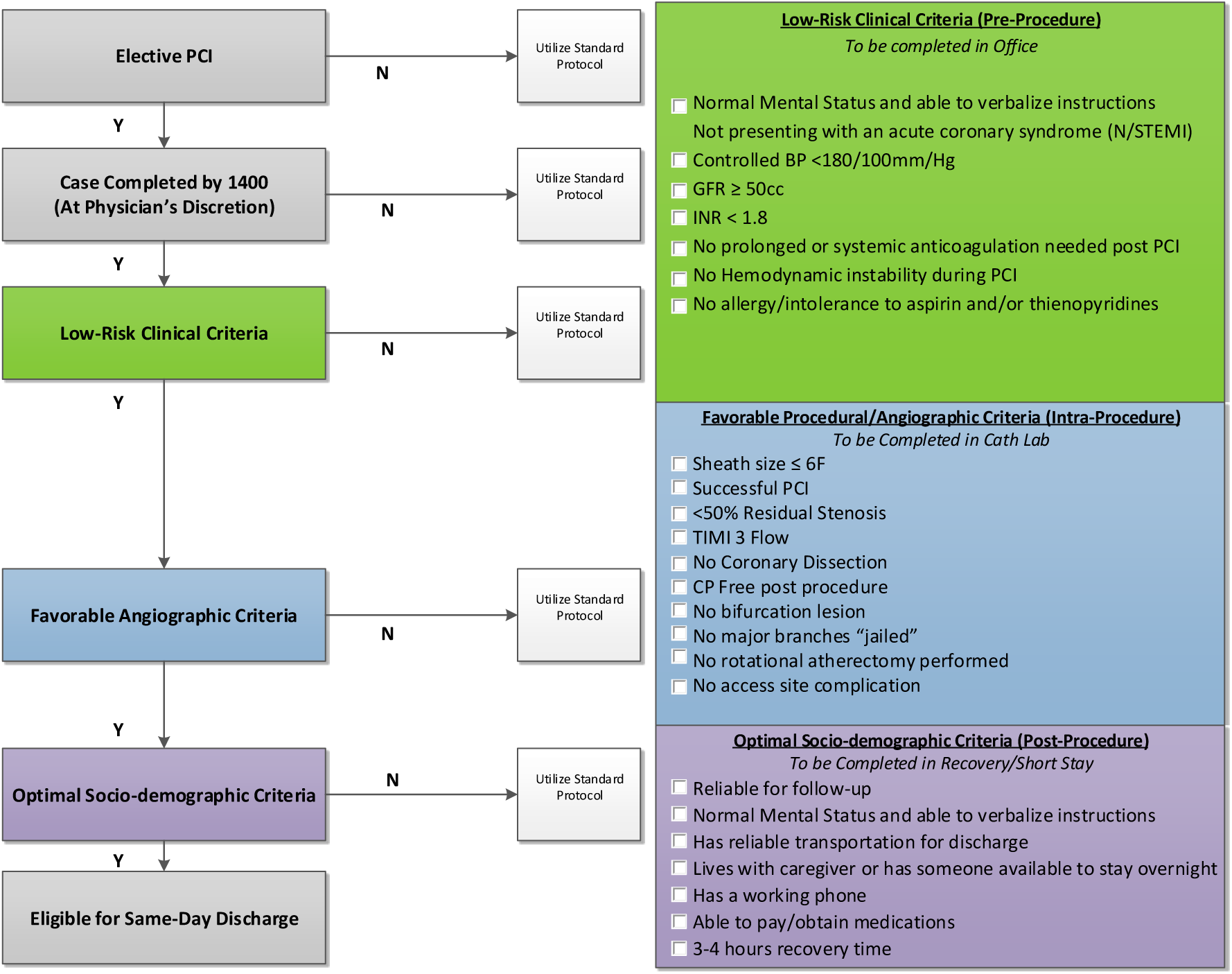
Same Day Discharge Protocol Diagram.

## References

1. Inohara T, Kohsaka S, Spertus JA, Masoudi FA, Rumsfeld JS, Kennedy KF, Wang TY, Yamaji K, Amano T, Nakamura M. Comparative Trends in Percutaneous Coronary Intervention in Japan and the United States, 2013 to 2017. J Am Coll Cardiol. 2020 Sep 15;76(11):1328–1340. doi: 10.1016/j.jacc.2020.07.037. PMID: 32912447.

2. Writing Group Members, Mozaffarian D, Benjamin EJ, et al. Heart Disease and Stroke Statistics-2016 Update: A Report From the American Heart Association. Circulation. 2016;133(4):e38–360. doi:10.1161/CIR.0000000000000350

3. Seto AH, Shroff A, Abu-Fadel M, et al. Length of stay following percutaneous coronary intervention: An expert consensus document update from the society for cardiovascular angiography and interventions. Catheter Cardiovasc Interv. 2018;92(4):717–731. doi:10.1002/ccd.27637

4. Venkitachalam L, Kip KE, Selzer F, Wilensky RL, Slater J, Mulukutla SR, Marroquin OC, Block PC, Williams DO, Kelsey SF; Investigators of NHLBI-Sponsored 1985-1986 PTCA and 1997-2006 Dynamic Registries. Twenty-year evolution of percutaneous coronary intervention and its impact on clinical outcomes: a report from the National Heart, Lung, and Blood Institute-sponsored, multicenter 1985-1986 PTCA and 1997-2006 Dynamic Registries. Circ Cardiovasc Interv. 2009 Feb;2(1):6–13. doi: 10.1161/CIRCINTERVENTIONS.108.825323. Epub 2008 Dec 15. PMID: 20031687; PMCID: PMC3024012.

5. Serruys PW, Rutherford JD. The Birth, and Evolution, of Percutaneous Coronary Interventions: A Conversation With Patrick Serruys, MD, PhD. Circulation. 2016 Jul 12;134(2):97–100. doi: 10.1161/CIRCULATIONAHA.116.023681. PMID: 27400895.

6. Bradley SM, Kaltenbach LA, Xiang K, Amin AP, Hess PL, Maddox TM, Poulose A, Brilakis ES, Sorajja P, Ho PM, Rao SV. Trends in Use and Outcomes of Same-Day Discharge Following Elective Percutaneous Coronary Intervention. JACC Cardiovasc Interv. 2021 Aug 9;14(15):1655–1666. doi: 10.1016/j.jcin.2021.05.043. PMID: 34353597.

7. Amin AP, Pinto D, House JA, et al. Association of Same-Day Discharge After Elective Percutaneous Coronary Intervention in the United States With Costs and Outcomes. JAMA Cardiol. 2018;3(11):1041–1049. doi:10.1001/jamacardio.2018.3029

8. Brayton KM, Patel VG, Stave C, de Lemos JA, Kumbhani DJ. Same-day discharge after percutaneous coronary intervention: a meta-analysis. J Am Coll Cardiol. 2013;62(4):275–285. doi:10.1016/j.jacc.2013.03.051

9. Bundhun PK, Soogund MZS, Huang WQ. Same Day Discharge versus Overnight Stay in the Hospital following Percutaneous Coronary Intervention in Patients with Stable Coronary Artery Disease: A Systematic Review and Meta-Analysis of Randomized Controlled Trials. PLOS ONE. 2017;12(1):e0169807. doi:10.1371/journal.pone.0169807

10. Kim M, Muntner P, Sharma S, et al. Assessing patient-reported outcomes and preferences for same-day discharge after percutaneous coronary intervention: results from a pilot randomized, controlled trial. Circ Cardiovasc Qual Outcomes. 2013;6(2):186–192. doi:10.1161/circoutcomes.111.000069

11. Madan M, Bagai A, Overgaard CB, et al. Same-Day Discharge After Elective Percutaneous Coronary Interventions in Ontario, Canada. J Am Heart Assoc. 2019;8(13):e012131. doi:10.1161/JAHA.119.012131

12. Taxiarchi P, Kontopantelis E, Martin GP, et al. Same-Day Discharge After Elective Percutaneous Coronary Intervention. Insights Br Cardiovasc Interv Soc. 2019;12(15):1479–1494. doi:10.1016/j.jcin.2019.03.030

13. Heyde GS, Koch KT, de Winter RJ, et al. Randomized Trial Comparing Same-Day Discharge With Overnight Hospital Stay After Percutaneous Coronary Intervention. Circulation. 2007;115(17):2299–2306. doi:10.1161/CIRCULATIONAHA.105.591495

14. Chambers CE, Dehmer GJ, Cox DA, et al. Defining the length of stay following percutaneous coronary intervention: an expert consensus document from the Society for Cardiovascular Angiography and Interventions. Endorsed by the American College of Cardiology Foundation. Catheter Cardiovasc Interv. 2009;73(7):847–858. doi:10.1002/ccd.22100

15. Rao, SV, Vidovich MI, Gilchrist IC, Gulati R, Gutierrez JA, Hess CN, Kaul P, Martinez S, Rymer J. 2021 ACC Expert Consensus Decision Pathway on Same-Day Discharge After Percutaneous Coronary Intervention: A Report of the American College of Cardiology Solution Set Oversight Committee. J Am Coll Cardiol. 2021 Feb, 77 (6) 811–825

16. Birger M, Kaldjian AS, Roth GA, Moran AE, Dieleman JL, Bellows BK. Spending on Cardiovascular Disease and Cardiovascular Risk Factors in the United States: 1996 to 2016. Circulation. 2021 Jul 27;144(4):271–282.

17. Collins CR, Wick E. Center for Medicare and Medicaid Services Bundled Payment for Care Improvement-Advanced for Major Bowel Surgery: Do We Have the Tools We Need to Succeed? J Am Coll Surg. 2019;229(4):S53. doi:10.1016/j.jamcollsurg.2019.08.129

18. Malenka DJ, Bhatt DL, Bradley SM, Shahian DM, Draoui J, Segawa CA, Koutras C, Abbott JD, Blankenship JC, Vincent R, Windle J, Tsai TT, Curtis J, Roe M, Masoudi FA. The National Cardiovascular Data Registry Data Quality Program 2020: JACC State-of-the-Art Review. J Am Coll Cardiol. 2022 May 3;79(17):1704–1712. doi: 10.1016/j.jacc.2022.02.034. PMID: 35483759.

19. Amin AP, Rao SV, Seto AH, Thangam M, Bach RG, Pancholy S, Gilchrist IC, Kaul P, Shah B, Cohen MG, Gluckman TJ, Bortnick A, DeVries JT, Kulkarni H, Masoudi FA. Transradial Access for High-Risk Percutaneous Coronary Intervention: Implications of the Risk-Treatment Paradox. Circ Cardiovasc Interv. 2021 Jul;14(7):e009328. doi: 10.1161/CIRCINTERVENTIONS.120.009328. Epub 2021 Jul 13. PMID: 34253050; PMCID: PMC8895384.

20. Lindner SM, McNeely CA, Amin AP. The Value of Transradial: Impact on Patient Satisfaction and Health Care Economics. Interv Cardiol Clin. 2020 Jan;9(1):107–115. doi: 10.1016/j.iccl.2019.08.004. PMID: 31733737; PMCID: PMC7772820.

21. Kirkpatrick JN, Hull SC, Fedson S, Mullen B, Goodlin SJ. Scarce-Resource Allocation and Patient Triage During the COVID-19 Pandemic: JACC Review Topic of the Week. J Am Coll Cardiol. 2020 Jul 7;76(1):85–92. doi: 10.1016/j.jacc.2020.05.006. Epub 2020 May 11. PMID: 32407772; PMCID: PMC7213960.

22. French G, Hulse M, Nguyen D, Sobotka K, Webster K, Corman J, Aboagye-Nyame B, Dion M, Johnson M, Zalinger B, Ewing M. Impact of Hospital Strain on Excess Deaths During the COVID-19 Pandemic – United States, July 2020-July 2021. MMWR Morb Mortal Wkly Rep. 2021 Nov 19;70(46):1613–1616. doi: 10.15585/mmwr.mm7046a5. PMID: 34793414; PMCID: PMC8601411.

